# Using electronic health record data to determine the safety of aqueous humor liquid biopsies for molecular analyses

**DOI:** 10.1101/2023.11.22.23298937

**Authors:** Julian Wolf, Teja Chemudupati, Aarushi Kumar, Joel A. Franco, Artis A. Montague, Charles C. Lin, Wen-Shin Lee, A. Caroline Fisher, Jeffrey L. Goldberg, Prithvi Mruthyunjaya, Robert T. Chang, Vinit B. Mahajan

**Author notes:** **Co-corresponding authors:** Vinit B. Mahajan M.D., Ph.D., Byers Eye Institute, Department of Ophthalmology, Stanford University, Palo Alto, CA, 94304 USA., Robert T Chang M.D., Byers Eye Institute, Department of Ophthalmology, Stanford University, Palo Alto, CA, 94304 USA. **Financial Support:** National Institutes of Health, Bethesda, Maryland, grant numbers: R01EY031952, R01EY031360, R01EY030151, and P30EY026877. The sponsor or funding organization had no role in the design or conduct of this research. **Address for reprints:**. **Précis:** This study demonstrates that aqueous humor liquid biopsies for molecular analyses can be obtained safely and at large-scale during intraocular surgery.

## Abstract

**Purpose:** Knowing the surgical safety of anterior chamber liquid biopsies will support the increased use of proteomics and other molecular analyses to better understand disease mechanisms and therapeutic responses in patients and clinical trials. Manual review of operative notes from different surgeons and procedures in electronic health records (EHR) is cumbersome, but free-text software tools could facilitate efficient searches.

**Design:** Retrospective case series.

**Participants:** 1418 aqueous humor (AH) liquid biopsies from patients undergoing intraocular surgery.

**Methods:** Natural language unstructured free-text EHR searches were performed using the Stanford Research Repository (STARR) cohort discovery tool to identify complications associated with anterior chamber paracentesis and subsequent endophthalmitis. Complications of the surgery unrelated to the biopsy were not reviewed.

**Main Outcome Measures:** Biopsy associated intraoperative complications and endophthalmitis.

**Results:** 1418 AH liquid biopsies were performed by 17 experienced surgeons. EHR free-text searches were 100% error-free for surgical complications, >99% for endophthalmitis (<1% false positive), and >93.6% for anesthesia type, requiring manual review for only a limited number of cases. More than 85% of cases were performed under local anesthesia without ocular muscle akinesia. Although the most common indication was cataract (50.1%), other diagnoses included glaucoma, diabetic retinopathy, uveitis, age-related macular degeneration, endophthalmitis, retinitis pigmentosa, and uveal melanoma. A 50-100μL sample was collected in all cases using either a 30-gauge needle or a blunt cannula via a paracentesis. The median follow-up was more than seven months. There was only one minor complication (0.07%) identified: a case of a small tear in Descemet’s membrane without long-term sequelae. No other complications occurred, including other corneal injuries, lens or iris trauma, hyphema, or suprachoroidal hemorrhage. There was no case of postoperative endophthalmitis.

**Conclusions:** Anterior chamber liquid biopsy during intraocular surgery is a safe procedure and may be considered for large-scale collection of AH samples for molecular analyses. Natural language free-text EHR searches are an efficient approach to reviewing intraoperative procedures.

## Introduction

Paracentesis of the anterior chamber (AC) has been used for decades to therapeutically lower intraocular pressure and remove pathological tissues such as blood or inflammatory cells. AC biopsies are an important diagnostic tool for patients with uveitis,^1^ and more recently, aqueous humor (AH) liquid biopsies are used for molecular testing in research. They allow the capture of locally enriched fluids containing thousands of molecules from healthy and diseased ocular tissues, that can be detected using a variety of molecular assays, including proteomics, metabolomics, and DNA sequencing. These tests offer the potential to understand disease mechanisms in living humans and to identify novel diagnostic and therapeutic strategies. Specifically, AH proteomics has been shown to predict the treatment response of patients with neovascular age-related macular degeneration and diabetic macular edema, two of the most frequent blinding diseases.^2, 3^ Analyzing cell-free DNA in AH has demonstrated potential to identify somatic genomic alterations in retinoblastoma, the most common eye cancer in childhood,^4, 5^ and can help to specify the underlying pathogen in patients with intraocular infections.^1, 6^ In addition, AH metabolomics studies have identified new potential biomarkers and therapeutic targets for diabetic retinopathy and glaucoma.^7, 8^ We recently found that state-of-the art techniques for increased AH proteomic throughput combined with single cell RNA sequencing of ocular tissues, allow cell level analyses in living patients, even in tissues like the retina that are not amenable for direct tissue biopsies.^9^

AH samples are routinely collected at the slit lamp in clinic mainly for diagnostic purposes and previous comparatively small studies have shown this to be a safe procedure in this setting.^10-13^ AC biopsies are also increasingly collected during human clinical trials to investigate target engagement and biomarkers associated with clinical outcomes.^14^ Large-scale collection of AH specimens can be performed at the beginning of intraocular procedures, including cataract, glaucoma, corneal, or vitreoretinal surgeries, which are among the most frequently performed surgeries worldwide. However, although part of a surgery, the AH biopsy may theoretically cause specific additional complications, such as corneal trauma, iris trauma, lens touch, hyphema, or suprachoroidal hemorrhage from hypotony.

Studies reviewing surgical procedures and their complications are challenging at large scale because coding in surgical reports frequently reflects only the general surgical procedures and not the complications. This requires tedious manual review of operative and postoperative notes, especially since different surgeons may use different language to describe intraoperative events. The requirement to manually read surgical reports can significantly limit the number of cases that can be reviewed. It has previously been shown that electronic health record (EHR) models that combine structured data with natural language free-text searches demonstrate improved accuracy.^15, 16^ Here, we used established EHR software tools and applied a natural language free-text searching strategy to assess complications in surgical reports and postoperative clinical notes. This allowed us to analyze the safety profile of AH biopsies at large scale, involving 1418 cases that were performed during intraocular surgery by many surgeons in a variety of cases. We found the procedure to be safe, creating new opportunities for molecular disease analysis in living humans.

## Methods

The study was approved by the Stanford University Institutional Review Board (IRB) and adhered to the tenets set forth in the Declaration of Helsinki. All patients underwent informed consent. Cases of intraocular surgery in which an AH liquid biopsy was performed at Stanford University between 2018 and 2023 were reviewed using the Stanford Research Repository (STARR) cohort discovery tool, Stanford Medicine’s research patient data repository for clinical and translational research.^17^ AH liquid biopsies were collected using a Mobile Operating Room Lab Interface (MORLI) that not only enables the immediate freezing of samples in the operating room, but is also linked to a REDCap (research electronic data capture) database ^18^ that allows point-of-care annotation of each sample with the patient identifiers (name, first name, date of birth, medical record number (MRN), procedure date) and other metadata including age, sex, and disease.^19, 20^ We searched the REDCap database for all intraocular surgery cases in which an AH liquid biopsy was performed and obtained the list of patient identifiers. To assess for intra- and postoperative complications associated to the AH biopsy, we then used the STARR tool to perform a natural language free-text search based on the operative reports and all clinical notes of each patient (**Figure 1**). The STARR database was queried using SQL code to 1) identify all study patients, 2) to confirm current patient identifiers and procedure dates, 3) to extract the full note(s) for review, and 4) to extract the anesthesia text from the note. All 17 surgeons were interviewed on the language they use in their operative reports and clinical notes to describe these complications. Based on their response, the following terms were used for the free-text search: “lens touch”, “iris touch”, “cornea touch”, “bleeding”, “hemorrhage”, “hyphema,”, “tear”, “leak”, “shallow”. “movement”, “suprachoroidal”, and “endophthalmitis”. Five-hundred operative and chart notes were manually reviewed to ensure accuracy of the search. Complications of the surgery that were not related to the AC biopsy were not tabulated for this study. The type of anesthesia was assessed in each case. We queried the STARR tool to obtain the operative report of each patient. We performed a free-text search in the operative reports using the following terms: “topical”, “intracameral”, “subtenon”, “peribulbar”, “retrobulbar”, “lidocaine”, and “general”. We then manually checked each case to ensure that the correct anesthesia type was selected, especially in those cases were more than 2 terms were found. We also identified the preoperative ophthalmic diagnoses and the primary surgical indication and determined the time of follow-up for each case.

**Figure 1:**
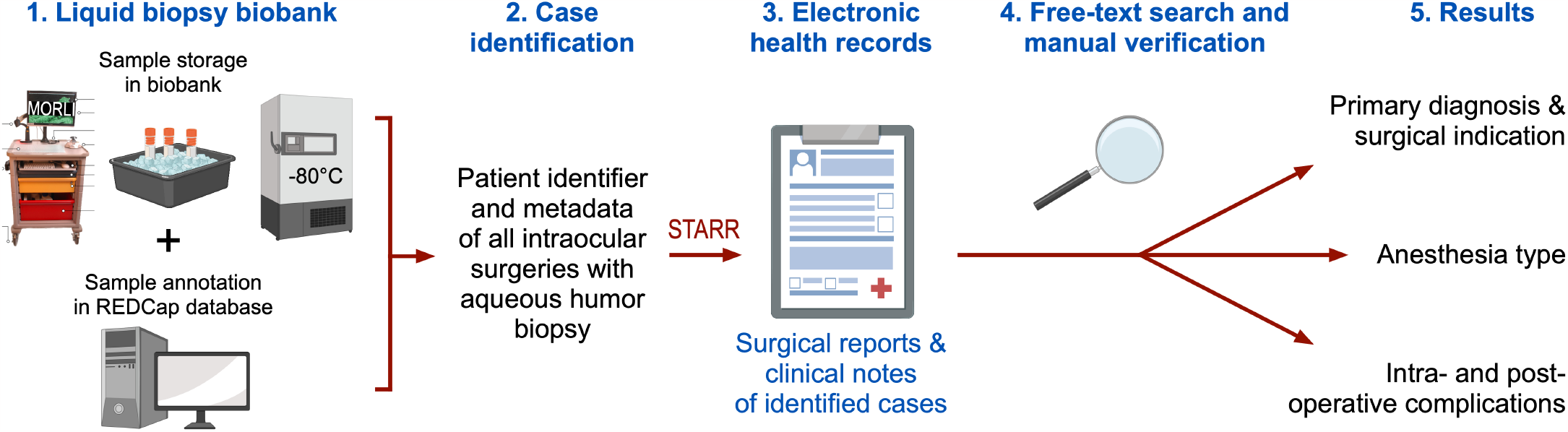
Electronic health record data analysis workflow. Our liquid biopsy biobank database was queried to identify patients that underwent intraocular surgery with aqueous humor liquid biopsy. The STARR tool was used to obtain electronic health records of these patients. We performed a natural language free-text search with manual verification to determine the primary diagnosis and surgical indication, the anesthesia type, and intra- and postoperative complications. MORLI: Mobile Operating Room Lab Interface, REDCap: research electronic data capture, STARR: Stanford Research Repository.

### Surgical technique

Anterior segment or vitreoretinal surgeries were performed with either topical anesthesia using eye drops, peribulbar anesthesia with a sub-Tenon’s injection, retrobulbar anesthesia under monitored anesthesia care, or general anesthesia. AH liquid biopsies were performed by 17 different ophthalmic surgeons and were all obtained using an operating microscope at the beginning of the surgery. Two techniques were followed based on surgeon preference (**Figure 2**). In the first technique, a surgeon inserted a 30-gauge needle connected to a 1 mL syringe perpendicular to the limbus and into the anterior chamber without prior incision. 50-100 μL of undiluted AH was manually aspirated, as previously described.^19^ In the second technique, a 15° blade was used to make a corneal incision perpendicular to the limbus in the superotemporal quadrant and an angled 30-gauge blunt cannula connected to a 1 mL syringe was inserted into the anterior chamber and 50-100 μL of AH was manually aspirated. In these cases, the corneal incision was part of the scheduled surgery. In both techniques, special attention was made to ensure the tip of the needle or the cannula remained over the peripheral iris in the mid anterior chamber to avoid damage to intraocular structures, including the corneal endothelium, iris, and lens. In vitrectomy cases, sclerotomies were created using a trocar-cannula system before the anterior chamber biopsy to ensure safe insertion of the trocars. After AH aspiration, the needle or the cannula were carefully removed from the AC and syringes were passed to the scrub technician. The fluid was expelled into a barcoded cryovial and the vial was immediately transferred on dry ice in the operating room and then prepared for storage in a biorepository at -80°C.^19^ The case continued as per required for the primary surgical indication.

**Figure 2:**
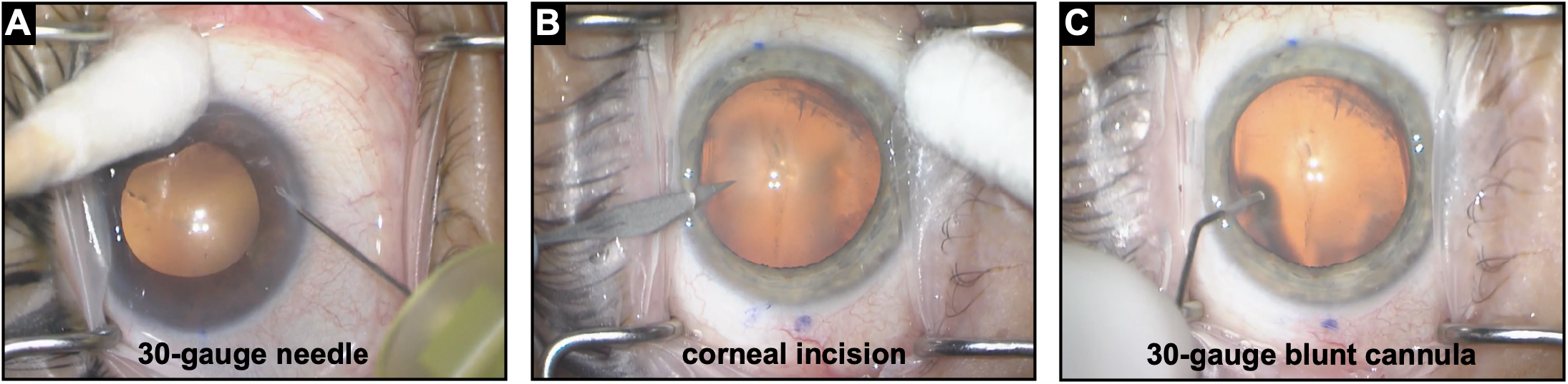
Two surgical techniques were applied to collect aqueous humor liquid biopsies during intraocular surgery. (**A**) In technique 1, a 30-gauge needle connected to a 1 mL syringe was inserted perpendicular to the limbus and into the anterior chamber without prior incision. (**B-C**) In technique 2, a 15° blade was used to make a corneal incision perpendicular to the limbus in the superotemporal quadrant (as part of the scheduled surgery) and an angled 30-gauge blunt cannula connected to a 1 mL syringe was inserted into the anterior chamber. In both techniques, 50-100 μL of undiluted AH was manually aspirated using the syringe.

## Results

The natural language free-text EHR search identified 1418 intraocular surgeries during which an AH liquid biopsy was performed. For 14 of these cases, there was a mismatch in the MRN, name, or date of birth fields between our REDCap database and the STARR EHR data (less than 1% of cases). We found this was due to spelling errors that occurred during data entry in the operating room in the REDCap database, such as a missing leading “0” in the MRN or a wrong date of birth. We were able to manually confirm the correct identity of each case using a combination of all identifiers and with reference to the STARR data. We manually examined the operative reports and the clinical notes to verify the results of the free-text search-based strategy and found no misclassification for intraoperative complications. The free-text search found 14 instances of “endophthalmitis” noted in patients EHRs. In all 14 instances, patients were seen for endophthalmitis prior to the AC tap, often years earlier, and there was no postoperative endophthalmitis following the AC tap. We also used a free-text search strategy in the operative reports to assess the type of anesthesia for each case. In 50 cases (3.5%), our search strategy did not find any hit, which was explained by the fact that none of the search terms about anesthesia type or only imprecise information (“monitored anesthesia care”) was provided in the anesthesia section of the surgical report. In those cases, we had to manually search the entire surgical report, but were able to determine the correct anesthesia type in each case. In 104 cases (7.3%), the term “general” was found using our free-text search, but our manual verification revealed that 40 of these cases (2.8%) were under local anesthesia combined with intravenous sedation. The reason for this discrepancy was that the term “general” was used imprecisely in the surgical report of these patients, most frequently as “transient general anesthesia”, although intravenous sedation was administered without any form of ventilation. We therefore manually classified these cases under the respective local anesthesia type. Overall, the natural language free-text EHR search strategy was 100% error-free for surgical complications (500 cases reviewed), >99% for endophthalmitis (<1% false positive), and >93.6% for anesthesia type, and despite the need for manual examination, greatly increased the efficiency of the search. In combination with manual annotation, we were able to obtain the intended information for each of the 1418 cases.

The 1418 AH liquid biopsies were performed on 1154 patients by 17 experienced surgeons with 6 surgeons collecting 50 or more biopsies each. More than 85 percent of the samples were collected under topical (1021), peribulbar (125), or subtenon (61) anesthesia. Only about 15 percent of the cases (211) were performed under ocular muscle akinesia using retrobulbar (147) or general (64) anesthesia. In 65.7 percent of cases, a 30-gauge needle was used without prior incision, while in the remaining 34.3 percent of cases, a blunt 30-gauge cannula was chosen, which was inserted following a corneal incision with a 15° blade. The most common primary surgical indications included cataract (711 specimens), cataract and glaucoma (207 specimens), glaucoma (147 specimens), retinal detachment (24 specimens), Fuchs’ endothelial dystrophy (21 specimens), and uveal melanoma (20 specimens). The remaining surgical indications are listed in **Table 1**. The intended volume of 50 – 100μL of undiluted AH was achieved in all cases.

**Table 1.**
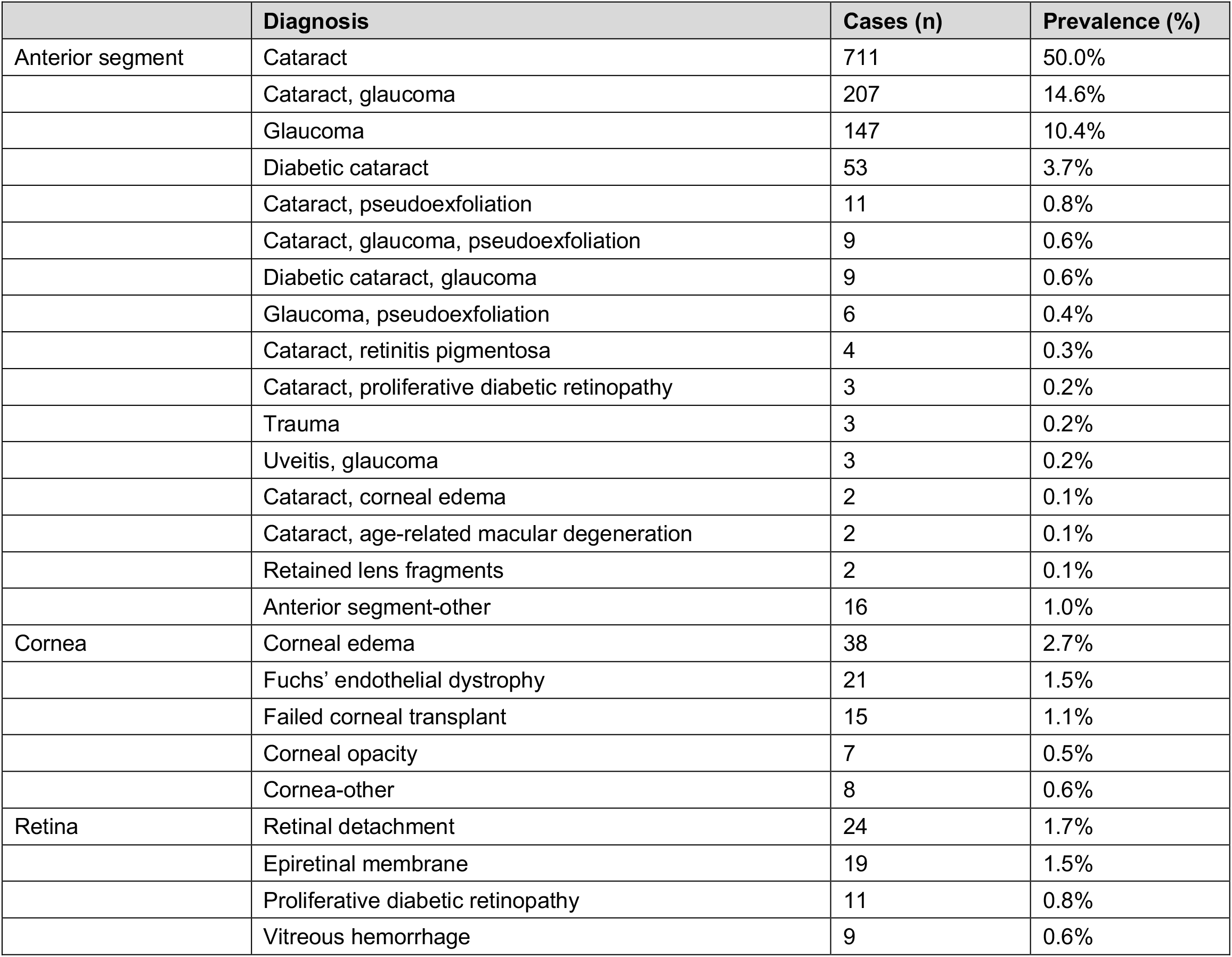
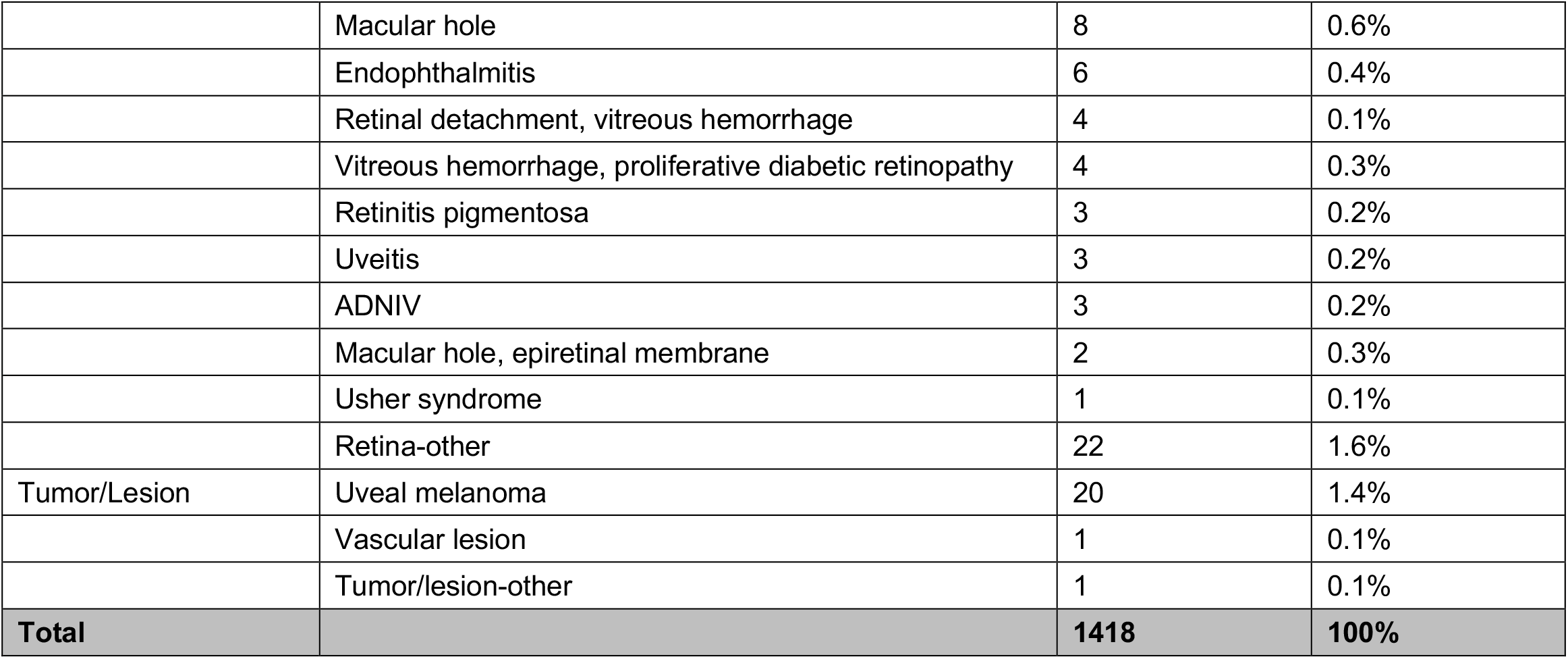
Aqueous humor liquid biopsy cases. A list of primary diagnoses for surgical cases in which an aqueous humor liquid biopsy was performed for proteomic analysis.

|  | Diagnosis | Cases (n) | Prevalence (%) |
| --- | --- | --- | --- |
| Anterior segment | Cataract | 711 | 50.0% |
|  | Cataract, glaucoma | 207 | 14.6% |
|  | Glaucoma | 147 | 10.4% |
|  | Diabetic cataract | 53 | 3.7% |
|  | Cataract, pseudoexfoliation | 11 | 0.8% |
|  | Cataract, glaucoma, pseudoexfoliation | 9 | 0.6% |
|  | Diabetic cataract, glaucoma | 9 | 0.6% |
|  | Glaucoma, pseudoexfoliation | 6 | 0.4% |
|  | Cataract, retinitis pigmentosa | 4 | 0.3% |
|  | Cataract, proliferative diabetic retinopathy | 3 | 0.2% |
|  | Trauma | 3 | 0.2% |
|  | Uveitis, glaucoma | 3 | 0.2% |
|  | Cataract, corneal edema | 2 | 0.1% |
|  | Cataract, age-related macular degeneration | 2 | 0.1% |
|  | Retained lens fragments | 2 | 0.1% |
|  | Anterior segment-other | 16 | 1.0% |
| Cornea | Corneal edema | 38 | 2.7% |
|  | Fuchs' endothelial dystrophy | 21 | 1.5% |
|  | Failed corneal transplant | 15 | 1.1% |
|  | Corneal opacity | 7 | 0.5% |
|  | Cornea-other | 8 | 0.6% |
| Retina | Retinal detachment | 24 | 1.7% |
|  | Epiretinal membrane | 19 | 1.5% |
|  | Proliferative diabetic retinopathy | 11 | 0.8% |
|  | Vitreous hemorrhage | 9 | 0.6% |
|  | Macular hole | 8 | 0.6% |
|  | Endophthalmitis | 6 | 0.4% |
|  | Retinal detachment, vitreous hemorrhage | 4 | 0.1% |
|  | Vitreous hemorrhage, proliferative diabetic retinopathy | 4 | 0.3% |
|  | Retinitis pigmentosa | 3 | 0.2% |
|  | Uveitis | 3 | 0.2% |
|  | ADNIV | 3 | 0.2% |
|  | Macular hole, epiretinal membrane | 2 | 0.3% |
|  | Usher syndrome | 1 | 0.1% |
|  | Retina-other | 22 | 1.6% |
| Tumor/Lesion | Uveal melanoma | 20 | 1.4% |
|  | Vascular lesion | 1 | 0.1% |
|  | Tumor/lesion-other | 1 | 0.1% |
| **Total** |  | **1418** | **100%** |

Detailed intraoperative complications were ascertained using the established Stanford Research Repository EHR search engines in combination with a natural language free-text search strategy.^17^ Out of 1418 biopsies, there was only one minor complication (0.07%) with no long-term sequelae (**Table 2**). In the case of a 70-year-old male patient with a 2+ cataract in the right eye undergoing cataract surgery under topical anesthesia, a 30-gauge cannula was used to obtain an AH specimen following a paracentesis made with a 15° super sharp blade. Upon AH withdrawal, there was shallowing of the AC. Balanced salt solution on a 30-gauge canula was used to reinflate the AC, at which time a small Descemet’s membrane tear was noted adjacent to the paracentesis. The remainder of the surgery was performed without incident, and the tear remained localized and small without extension. Upon postoperative follow up over 3 months, there was no residual corneal edema or other sequelae associated with this tear. None of the other patients had any trauma to the lens or iris. No other complications, such as hyphema, entry site leak, anterior chamber shallowing forcing termination of fluid collection, suprachoroidal hemorrhage, or problems caused by patient’s movements were noted.

**Table 2.** Incidence of complications with aqueous humor liquid biopsies (n = 1418).

| Complication | Number (%) |
| --- | --- |
| Endophthalmitis | 0 (0%) |
| Lens touch/trauma | 0 (0%) |
| Iris touch/trauma | 0 (0%) |
| Hyphema | 0 (0%) |
| Descemet's membrane tear | 1 (0.07%) |
| Other corneal trauma | 0 (0%) |
| Entry site leak | 0 (0%) |
| Anterior chamber shallowing forcing termination of fluid collection | 0 (0%) |
| Suprachoroidal hemorrhage | 0 (0%) |
| Problems caused by patient's movements | 0 (0%) |
| **Total** | **1 of 1418 (0.07%)** |

The median follow-up time at our institution was more than seven months (223 days, range: 0 – 2003 days). Ninety-nine percent (1408) of the cases were seen for an immediate follow-up the next day; 97.1% (1377), 88.6% (1256), and 69.0% (978) were followed-up for one week, one month, and three months, respectively. There were no cases of endophthalmitis following the AH biopsy.

## Discussion

In this study, we reviewed a large cohort of intraocular surgeries in which an AH liquid biopsy was performed regardless of the primary surgical indication and found only one minor complication related to obtaining the AH specimen, corresponding to a complication rate of 0.07%. Importantly, we did not observe any case of intraocular inflammation or endophthalmitis at for all patients with one week follow-up (97.1%) and for all patients that had a 3-month post biopsy follow-up (69.0%). These findings indicate that obtaining an AH biopsy during intraocular surgery using a 30-gauge needle or cannula is a safe procedure. Our study further demonstrates that EHR analyses that incorporate natural language free-text searches are helpful to investigate intra- and postoperative complications, especially when examining operative reports that contain data not reflected in billing codes or “big-data” clinical repositories.

In smaller studies with mainly uveitis patients, AH biopsy was reported as a safe procedure when performed at the slit lamp in the outpatient clinic. Van der Lelij et al. investigated 361 AH biopsies obtained after preincision with a blade for diagnostic assessment in patients with uveitis.^10^ Apart from a small hyphema in 5 of 72 cases (6.9%), no other complications were reported. The largest study so far investigated 560 AH biopsies in patients with uveitis performed at the slit lamp without preincision and found 4 complications (0.7%), among them 1 case of a lens touch, 2 cases of air in the AC, and 1 case of an allergic reaction to the disinfectant.^12^ Since air in the AC is self-limiting and typically does not have significant consequences, the complication rate could be considered to be 1 out of 560 cases (0.2%). Two other similar studies reported air in the AC in 2 out of 70 cases (2.9%) ^11^ and no complication in 301 cases.^13^ Our findings in a cohort more than 2.5 times larger demonstrate that AH liquid biopsies can be safely obtained in the operating room across a variety of intraocular surgeries, ranging from corneal interventions to cataract and glaucoma surgery, and to vitreoretinal procedures. One of the potential challenges of collecting AH samples at the slit lamp is that the patient can move the eye. During intraocular surgery in the operating room, retrobulbar and general anesthesia make the eye immobile, whereas under local anesthesia, the patient could still move the eye. In our study, more than 85 percent of the cases were under local anesthesia without ocular muscle akinesia, but no complications due to eye movements were observed. These findings suggest AC biopsies in an outpatient procedure room with an operating microscope and supine patient might reduce some complications reported when using an exam lane slit lamp with an upright patient. In addition, compared to AC taps performed as a stand-alone procedure at the slit lamp, sampling AH during intraocular surgery has the advantage that the risks generally do not exceed the risks of already planned intraocular surgery. Collecting these samples on a large scale could significantly enhance molecular analyses, including genomic, proteomic, and metabolomic studies, with the goal to improve our understanding of disease mechanisms in living humans.^1-6, 8, 9^

In this study, two different surgical techniques were followed, one using a 30-gauge needle without prior incision and another using a blunt 30-gauge cannula that was inserted following a corneal incision with a blade. Using a 30-gauge needle has the advantage that the AC may be less likely to shallow, the wound is self-sealing, and the risk of blood contamination of the sample is minimized. Inserting a blunt cannula through the prior corneal incision that is generated as part of many intraocular surgeries has the advantage that no additional entry site must be created and that the risk of damage to intraocular structures may be lower compared to the first technique. Instead of the blunt cannula, a sharp 30-gauge needle may also be inserted through the corneal incision. However, with the two steps, the AC may be more likely to leak some fluid and shallow, thus the volume that can be collected could be lower. A single minor complication we observed was a small tear in Descemet’s membrane following insertion of the 30-gauge blunt cannula, but there was no serious impact on the rest of the case or on postoperative healing. Several insertions of the blunt cannula were also part of the normal surgery to deliver balanced salt solution and viscoelastic. Taken altogether, the results suggest that both techniques are safe and surgeon preference can be followed.

There are limitations to this study. We performed a multi-instant search that included various EHR data for each patient, including the operative report and all clinical notes. Free-text searches are subject to spelling errors, but even if a term to describe a complication was misspelled once, it’s very unlikely that it was misspelled consistently in all documents that were searched. Importantly, our manual review confirmed that no complications were systematically missed. The anesthesia data had the highest inaccuracy. The reason was inaccurate use of terms to describe anesthesia type in the surgical report. Interviewing the anesthesia team to learn how the terms are used and applying more specific search terms may improve the accuracy in future studies. Follow-up for some cases was not available beyond three months, and it seems unlikely that the AH biopsy prior to a complete intraocular surgery would trigger complications beginning after three months.

Nonetheless, a prospective, randomized controlled trial analyzing all postoperative complications between cases with or without an AH biopsy would need to be conducted. Alternatively, a big data approach that reviewed EHR of tens of thousands of surgery cases might reveal an association with complications.

In conclusion, in this large cohort of 1418 AH liquid biopsies that were collected during intraocular surgeries demonstrated less than a 0.07% minor complication rate related to the sample collection itself. Our findings indicate that obtaining AH specimens during intraocular surgery is a safe procedure. Collecting these samples for research or in clinical trials could significantly improve molecular studies leading to a better understanding of disease pathophysiology in living humans. Our results further demonstrate that implementing natural language free-text search in reviewing EHR could increase the size and quality of retrospective studies assessing surgical complications.

## Data Availability

All data produced in the present study are available upon reasonable request to the corresponding authors.

## Acknowledgements

This research used data or services provided by STARR, “STAnford medicine Research data Repository,” a clinical data warehouse containing live Epic data from Stanford Health Care, the Stanford Children’s Hospital, the University Healthcare Alliance and Packard Children’s Health Alliance clinics and other auxiliary data from Hospital applications such as radiology PACS. STARR platform is developed and operated by Stanford Medicine Research Technology team and is made possible by Stanford School of Medicine Research Office. Selected illustrations were created with BioRender.com.

## Notes

**Conflict of Interest:** No conflicting relationship exists for any author. Running head: Safety of aqueous biopsies during intraocular surgery

### Competing Interest Statement

The authors have declared no competing interest.

### Funding Statement

This study was funded by the National Institutes of Health, Bethesda, Maryland, grant numbers: R01EY031952, R01EY031360, R01EY030151, and P30EY026877. The sponsor or funding organization had no role in the design or conduct of this research.

### Author Declarations

The Institutional Review Board (IRB) of Stanford University gave ethical approval for this work.

